# A multicentre, prospective, randomised controlled trial to evaluate hexaminolevulinate photodynamic therapy (Cevira^®^) as a novel treatment in patients with high grade squamous intraepithelial lesion: APRICITY Phase 3 study protocol

**DOI:** 10.1101/2021.12.02.21266991

**Authors:** Fei Chen, Zoltán Novák, Christian Dannecker, Long Sui, Youzhong Zhang, Zhixue You, Ling Han, Jinghe Lang, Peter Hillemanns

## Abstract

**Introduction:** High-risk human papilloma virus (HPV)-associated cervical cancer is the fourth most common cancer in women worldwide. Current treatments of high grade squamous intraepithelial lesion (HSIL) of the cervix are based on invasive surgical interventions, compromising cervical competence and functionality.

**Objective:** APRICITY is a multicentre, prospective, double-blind, randomised controlled Phase 3 study further evaluating the efficacy and safety of Cevira^®^, an integrated drug- and light-delivery device for hexaminolevulinate photodynamic therapy, which shows promise as a novel, non-invasive therapy for women with HSIL.

**Methods and analysis:** Patients with biopsy-confirmed HSIL histology are invited to participate in the study currently being conducted at 47 sites in China and 25 sites in Ukraine, Russia and European Union. The aim is to include at least 384 patients which will be randomised to either Cevira^®^ or placebo group (2:1). All patients will be assessed 3 months after first treatment and a second treatment will be administered in patients who are HPV positive or have at least low grade squamous intraepithelial lesion (LSIL). Primary endpoint is the proportion of the responders at 6 months after first treatment. Secondary efficacy endpoints and safety endpoints will be assessed at 6 months, and data for secondary performance endpoints for Cevira^®^ device will be collected at 3 months and 6 months, in case second treatment was administered. All patients in the Cevira^®^ group will be enrolled in an open, long-term extension study following patients for further 6 months to collect additional efficacy and safety data (study extension endpoints).

**Conclusion:** Due to its non-invasiveness and convenient application, Cevira^®^ may be a favourable alternative to surgical methods in treatment of patients with HSIL.

**Ethics and dissemination:** The study was approved by the ethics committee of the Peking Union Medical College Hospital and Hannover Medical University, Germany. Findings will be disseminated through peer review publications and conference presentations.

**Trial registration number:** clinicaltrials.gov NCT04484415

## Introduction

Cervical cancer is the fourth most common cancer in women worldwide, almost exclusively linked to infection with high-risk human papillomaviruses (hrHPV) (1). In 2020, 604,000 new cases and 342,000 deaths worldwide were attributed to cervical cancer (2). About 85% of new cases are occurring in developing regions with high HPV prevalence such as Asia, Africa and Eastern Europe (HPV prevalence of 45.5%, 29.6% and 21.4%, respectively (3, 4). In China, the most prevalent HPV subtypes are HPV16, 52 and 58, while in Europe HPV16, 31 and 33 are the most common subtypes (5). Notably, all mentioned HPV subtypes belong to the same alpha genotype (6). There are at least 14 high-risk HPV subtypes identified, with HPV16 and HPV18 causing 70% of cervical cancers and pre-cancerous lesions (5, 7).

HPV is transmitted during sexual intercourse with the highest prevalence among sexually active young women. In the vast majority (∼90%) infection is spontaneously cleared and induced low grade squamous epithelial lesion (LSIL) or cervical intraepithelial neoplasia (CIN) 1 has low potential to develop into cervical malignancy. Nevertheless, a subset of patients is at risk to develop persistent HPV infection increasing the risk for progression to high grade squamous intraepithelial lesion (HSIL) and eventually cancer (8, 9). Based on histopathological characteristics and the severity of dysplasia, HSIL can be subdivided into cervical intraepithelial neoplasia (CIN) 2 and 3, corresponding to moderate and severe dysplasia, respectively (10, 11). Unlike LSIL which usually resolves spontaneously, HSIL mostly requires medical treatment.

Current treatment options for patients with HSIL include excisional and ablative treatment (12). However, these surgical treatments may lead to perinatal complications, including preterm labour, low birth weight and perinatal death, limiting their use in women of reproductive age (13). Surgical treatments lead to a success rate of 85-95% in complete excision of the lesion (14). Recurrences occur as precancerous conditions such as CIN2 or CIN3, however, there is an elevated risk for invasive cervical cancer as well (15, 16). To preserve cervical tissue functionality, repeated surgical interventions are not recommended.

Non-invasive therapies have been developed for the treatment of HSIL and include topical agents (immune-modulators, anti-proliferative medications, antivirals, herbal regimens and probiotics), therapeutic vaccines and biologicals (17-19). However, due to the lack of sufficient clinical evidence, none of them have been accepted by the American Society for Colposcopy and Cervical Pathology (ASCCP) and European Federation for Colposcopy (EFC) for the management of cervical cancer and precancerous lesions and surgical methods remain the standard of care (12, 20).

Due to the side-effects associated with surgical treatments and the lack of evidence for most of the current non-invasive therapies, there has been a growing interest in non-invasive photodynamic therapy (PDT) using topically applied photosensitizers for the treatment of CIN (21-26). PDT is based on the accumulation of a photosensitizer or its precursor in the target cells, which upon illumination generates reactive oxygen species (ROS) that eradicate the diseased cells by inducing apoptosis and necrosis while preserving the underlying stroma and thereby the functionality of the cervix (27). For the treatment of CIN, topical hexaminolevulinate hydrochloride (HAL) has been mostly studied as photosensitizer showing promising efficacy and favourable safety results (24, 25). These initial results were confirmed in a Phase 2b study administrating HAL as an ointment via an intravaginal photoactivation device (Cevira^®^, Photocure ASA, Oslo, Norway) (26).

The objective of APRICITY Phase 3 multicentre, prospective, randomised controlled trial (RCT) is to further evaluate the efficacy and safety of Cevira^®^ compared to placebo in the treatment of patients with cervical histological HSIL (i.e. CIN2/3).

## Methods and analysis

### 2.1 Study design

The Phase 3 study is designed as a multicentre, prospective, double-blind RCT enrolling patients with an adequate colposcopy and histology diagnosis of HSIL (clinicaltrials.gov Identifier: NCT04484415) (Figure 1). Randomisation to either Cevira^®^ or placebo (2:1) is stratified by CIN diagnosis (CIN2 or CIN3) and HPV status (HPV-, HPV16+ or HPV18+Other+). Primary efficacy will be evaluated 6 months after first treatment for both groups. A second treatment will be administered in patients from both treatment groups who at the 3-month assessment have cytology of LSIL or more severe lesion (HSIL or atypical squamous cells-cannot exclude HSIL [ASC-H]) or in patients who are HPV positive. Retreatment visit should be no later than 1 month after the 3-month assessment visit.

**Figure 1.**
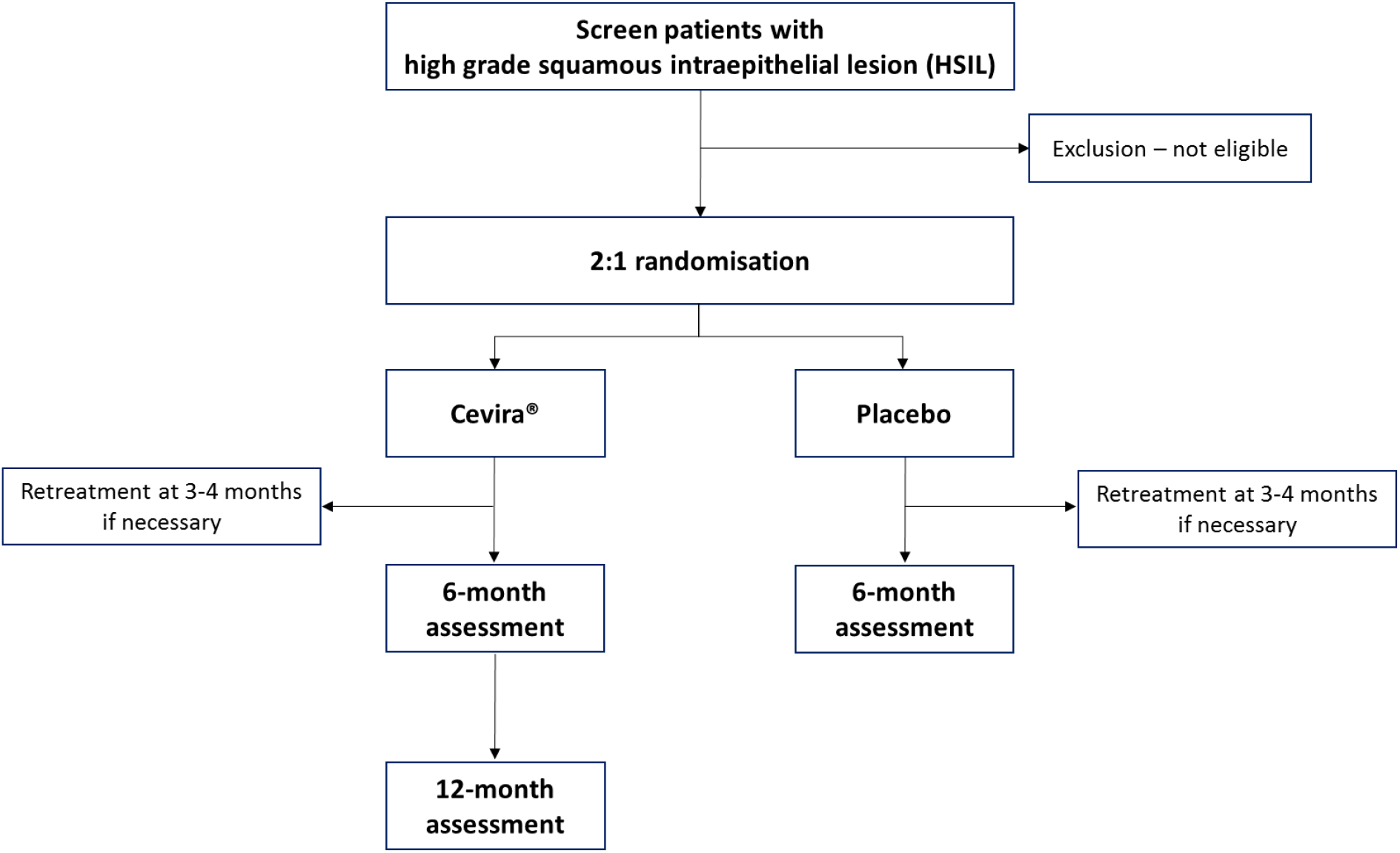
Overview of the study design.

All patients in the Cevira^®^ group will be enrolled in an open, long-term extension study following patients for an additional 6 months. To minimize the risk to the patients in the placebo group, they will be unblinded after 6 months to complete the study.

The study will be conducted at 47 sites in China and 25 sites in Ukraine, Russia and the European Union (EU, including Hungary, Romania, Germany, Czech Republic, Slovakia, Poland and Netherlands).

### 2.2 Study population

In Phase 2b study, the efficacy of HAL PDT could be demonstrated only in CIN2 patients, probably due to the high spontaneous regression rate in the CIN1 population (26). Based on this outcome, it was decided to only enrol patients with HSIL (CIN2/3) in the present Phase 3 study.

### 2.3 Eligibility criteria

#### 2.3.1 Inclusion criteria

To be included, patients must have biopsy-confirmed HSIL histology determined by a panel of three pathologists from a central laboratory in each region (China, US and Europe), not more than 2 months prior to the administration of Cevira^®^ or placebo. Colposcopy should visualise the entire lesion margin and entire cervical transformation zone, including the squamocolumnar junction, to demonstrate that the lesion covers more than 15% of the uterine cervix before biopsy. Additionally, the uterine cervix should have an average diameter of approximately 27 mm to allow application of Cevira^®^. Only female patients aged 18 years and older will be included. Patients must use a highly effective method of contraception during the entire study and 30 days after study end. Sterilised women or women who are post-menopausal for at least 1 year can be included without use of contraception.

#### 2.3.2 Exclusion criteria

Key exclusion criteria are a total lesion area covering over 50% of the cervix (only for biopsy-confirmed CIN3), invasive cervical cancer, adenocarcinoma in situ or other glandular intraepithelial lesions and lesions extending to the cervical canal or vaginal vault. Of note, in certain countries (e.g. Hungary), the ethical review board allowed only the inclusion of HSIL/CIN2 patients and excluded patients diagnosed with CIN3. Additional exclusion criteria are significant vaginal infection or bleeding and porphyria. Furthermore, patients must not be pregnant or breastfeeding.

Prior and during the entire study follow-up, patients are not allowed to use drugs or treatments that may affect efficacy evaluation, i.e., drugs treating HPV, HSIL and tumours as well as regulating immunologic function.

### 2.4 Interventions

Cevira^®^ is an integrated combination of 5% HAL in ointment and the drug delivery device Cevira^®^ CL7 (Figure 2A). The drug is administered intravaginally to the cervix by a gynaecologist using the drug delivery device (Figure 2B). The device is a single-use, disposable, LED-based integrated red light source used to photoactivate the drug. The device will automatically switch on the light 5 hours after administration and provide continuous photoactivation of 125 J/cm^2^ over 4.6 hours before automatically shutting down. The device needs to be removed by the patient once the treatment has been completed between 11 to 24 hours after administration. The placebo ointment contains only vehicle and is similar in appearance and consistence to the Cevira^®^ ointment. The placebo device is identical in appearance as the Cevira^®^ CL7 device without providing light.

**Figure 2:**
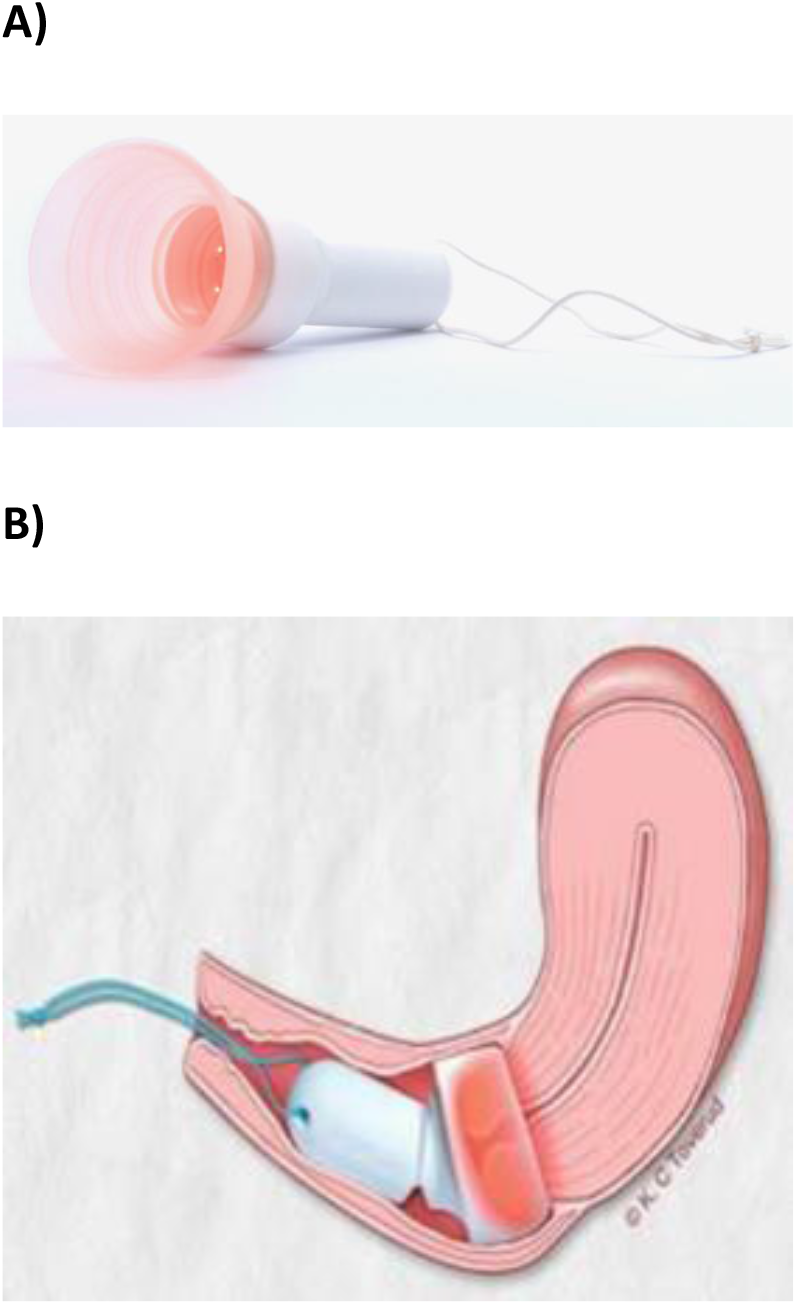
Cevira^®^ (Photocure ASA, Oslo, Norway). A) Cevira^®^ CL7 drug delivery device with integrated red light source. B) Ointment and device are administered intravaginally by the gynaecologist to the cervix of the HSIL patient.

### 2.5 Outcomes

The primary endpoint is the proportion of the responders at 6 months after the first treatment, with response being defined as normal histopathology or LSIL histopathology with clearance of baseline HPV. The list of secondary efficacy endpoints, study extension endpoints and safety endpoints can be found in Table 1.

**Table 1:** Secondary efficacy endpoints, study extension endpoints and safety endpoints of APRICITY Phase 3 study of Cevira^®^.

|  |
| --- |
| **<u>Secondary efficacy endpoints</u>** |
| a) The proportion of HPV(+) patients with clearance of baseline HPV at 6 months after the first treatment<br>b) The proportion of HPV16(+) patients with clearance of HPV16 at 6 months after the first treatment<br>c) The proportion of HPV16 and/or HPV18(+) patients with clearance of baseline HPV at 6 months after the first treatment<br>d) The proportion of patients with histologic regression, defined as LSIL or normal histology, at 6 months after the first treatment |
| **<u>Secondary performance/usability endpoints</u>** |
| a) The proportion of gynaecologists successfully inserting the device within 15 minutes<br>b) The proportion of patients with device dislocation or slippage during treatment<br>c) The proportion of patients removing the device outside the specified time |
| **<u>Study extension endpoints</u>** |
| a) The proportion of patients who had LSIL histology and non-clearance of baseline HPV at 6 months, who became responders at 12 months after the first treatment<br>b) The proportion of responders at 6 months who have continued regression at 12 months after the first treatment |
| **<u>Safety endpoints</u>** |
| a) The proportion of patients with AEs up to 6 months after the first treatment<br>b) The proportion of patients with Cevira® 5% HAL ointment-related AEs up to 6 months after the first treatment<br>c) The proportion of patients with Cevira® CL7 device-related AEs up to 6 months after the first treatment<br>d) The proportion of device deficiencies<br>e) The proportion of patients in the treatment group with AEs up to 12 months after the first treatment |

### 2.6 Assessments

The primary and secondary efficacy assessments will be done 6 months after first treatment and will be based on histology and clinically validated testing for HPV (Cobas, Roche). Colposcopy-directed biopsies will be obtained from all colposcopically-suspicious areas. If there is a normal colposcopy at 6 months or at assessment of study extension endpoints at 12 months, biopsies will be obtained from the original baseline affected area(s) to confirm histologic regression. Two pathologists will independently review the slide(s) from each biopsy in a blinded manner. If there is a discrepancy between the biopsy diagnosis, a third pathologist will review the slide(s). Clinically validated testing of cytology (ThinPrep, Hologic) and HPV (Cobas, Roche) will be used to determine the need for a second treatment.

The secondary performance assessment will be based on the number of gynaecologists successfully inserting the device within 15 minutes, the number of patients with device dislocation or slippage during treatment and the number of patients removing the device outside the specified time. Data will be collected using a patient diary and assessed at 3 months and at 6 months, in case a second treatment was applied. Safety endpoints assessment will be done at 6 months after first treatment.

### 2.7 Statistical considerations

#### 2.7.1 Sample size

The sample size calculation is based on the efficacy results for the HSIL histology population from the Phase 2b study using a significance level of 5% (26). It is assumed that the proportion of patients who will achieve response is 60% in the Cevira^®^ group and 40% in the placebo group. To detect this difference with 90% power, 209 patients need to be included in the Cevira^®^ group and 105 patients in the placebo group using a 2:1 randomisation.

Based on the Phase 2b study results, an 8% error rate of pathological assessment and 10% dropout rate should be considered. Therefore, the total sample size needs to consist of at least 384 patients (256 in Cevira^®^ group and 128 in the placebo group). The aim is to enrol 300 patients in Chinese centres and 84 patients in centres in EU, Ukraine and Russia, with no recruitment goal for the sites. Patients, where the diagnosis for study enrolment was changed from HSIL to not HSIL, will be excluded from the primary efficacy analysis (modified intent-to-treat population, mITT).

#### 2.7.2 Statistical analysis plan

To avoid bias, the statistical analysis team will be required to remain blinded throughout the entire study period until primary database lock (primary analyses will be performed when all patients have either completed the 6 month assessments or are early terminated from the study).

The primary efficacy endpoint analysis will be performed on the mITT population and repeated on per protocol (PP) population, which is defined as the subset of patients in the mITT population who had no major protocol violations. Analysis will be done using the Cochran Mantel Haenszel (CMH) test stratified by CIN diagnosis (CIN2 or CIN3) and HPV status (HPV-, HPV16+ and HPV18+Other+). Estimates and exact 95% confidence intervals for the proportion of patients who achieve response will be calculated overall, for each treatment group, for each diagnosis group, for each HPV status group and for each diagnosis group by HPV status subgroup (i.e. for each of the randomization strata).

The secondary efficacy endpoints on mITT population will be analysed the same as described for the primary endpoint. The study extension endpoints will be summarised using counts and percentages for the extension population. The summary will be presented overall, by CIN diagnosis and by HPV status. The safety analysis will be performed on the safety population. If possible, a distinction will be made between Cevira^®^ 5% HAL ointment-related and Cevira^®^ CL7 device-related AEs.

### 2.8 Randomisation and blinding method

The patients will be randomised through an Interactive Web Response System (IWRS) after initial screening by the investigators. The IWRS will generate a randomisation number after the investigator inputs the required information and will then assign a product for the patient. The investigators, study personnel and patients are blinded to the treatment groups as Cevira^®^ and placebo products are identical in packaging. Additionally, the light signal before insertion does not differ between Cevira^®^ and placebo products. If during the blinded part of the study a medical emergency or SAE occurs and the patient’s condition requires knowledge of the test medication, the study blind may be broken and reported for that specific patient. After the 6-month assessment, the planned unblinding procedure will be performed by the investigators to decide if patients will continue in the open-label extension study.

### 2.9 Data and study monitoring

An electronic data collection (EDC) system will be used to collect and manage the trial data in this study. Patient data should be entered continuously during the study and within 48 hours after a visit is performed. Study monitoring will be performed in accordance with International Conference on Harmonization (ICH) E6-Good Clinical Practice (GCP)/ISO 14155:2020 as applicable, the sponsor/contract research organization (CRO) standard operating procedures (SOPs), the protocol, the monitoring plan and applicable local regulations. If missing or inconsistent data not catered for are detected, queries will be issued. Queries may also be generated during the data validation process and shall be resolved immediately before database lock. All study documentation at the investigator site and sponsor site will be archived in accordance with the ICH E6-GCP/ISO 14155:2020 as applicable, EU Regulation 536/2014, 21 CFR 312.62, and the sponsor’s quality standards and SOPs. An auditor authorised by the sponsor may audit the investigational site and request access to all source documents, electronic case report form (eCRF) and other study documentation.

### 2.10 Ethical considerations

The study was approved by the ethics committee of the Peking Union Medical College Hospital on 2^nd^ of July, 2020 (Nr. KS20202255). It is conducted in accordance with the International Conference on Harmonization Guideline for Good Clinical Practice (ICH-GCP), the Declaration of Helsinki and all applicable national and international laws, regulations and standards, including archiving of essential documents. Patients agreeing to participate in the study must sign an informed consent form approved according to local regulations. The study site staff member conducting the consent process must also sign the consent form on the same occasion. All amendments to the clinical study protocol should be agreed between the sponsor and the investigator and be recorded with a justification for the amendment. The only exceptions are where necessary to eliminate an immediate hazard to study patients, or when the changes involve only administrative aspects of the study (e.g. change in monitor(s), change of telephone number(s)). All information concerning drug/device and the sponsor’s research and product development is considered confidential and will remain the sole property of the sponsor. A financial agreement will be signed between the sponsor and the investigators and/or the institution involved as required.

## Results

APRICITY Phase 3 study of Cevira^®^ is currently recruiting patients in Europe and China. Recruitment started in November 2020 with the aim to enrol at least 384 patients over the period of 15 months. Planned overall duration of the study is 34 months, with last patient last visit planned in September 2023.

As of November 12, 2021, 188 patients have been enrolled in China and 6 patients in Europe, with 60% and 40% of patients being diagnosed with CIN2 with CIN3, respectively. HPV status of the enrolled patients: HPV-(3%), HPV16+ (59%) and HPV18+Other+ (38%). The majority of patients was aged 21-40 years (93%), with 1% of younger women (age group 18-20 years) and 4.3% of women over 40 years of age.

As of 1^st^ of November 2021, 54 sites have been initiated with 45 sites in China, 3 in Hungary, 2 in Ukraine, 2 in Germany and 2 site in Slovakia. In total, 194 patients have been enrolled.

## Discussion

Cervical cancer is the most common HPV-related malignancy (7). Given the substantial burden of cervical cancer globally, efforts have been made to develop effective prevention measures, including HPV vaccination and screening programmes in combination with timely and efficient treatment of pre-cancerous lesions (2, 11). Unfortunately, these preventive measures are not equally implemented worldwide, with absent or inadequate screening and vaccination programmes in many low- and middle-income countries who suffer the highest HPV incidence rates. Especially in Eastern Europe and Central Asia, a rapid increase in premature cervical cancer mortality has been reported in recent generations (2). Furthermore, currently available vaccines are expensive and directed against only certain HPV subtypes (4, 11).

PDT has been clinically approved for the treatment of different cancers, including skin cancer, superficial oesophageal cancer and lung cancer (28-30). Due to the lack of adequate non-surgical treatment modalities, the potential of PDT for the treatment of CIN has been investigated in this and previous studies (21-26, 31). The main advantage of PDT for the treatment of CIN is its non-invasiveness, leaving the cervix intact and thereby preserving fertility. Moreover, targeted PDT of CIN is not restricted by HPV subtype causing pre-cancerous lesions.

For the treatment of CIN, topical administration of both 5-aminolaevulinic acid (5-ALA) and its esterified derivate HAL have been studied, with HAL being preferred due to its better stability and increased fluorescence at lower doses leading to less systemic exposure (21, 23, 28). Topically intravaginal administrated HAL is selectively taken up by epithelial HSIL cells upon which it is metabolised into the photosensitizer protoporphyrin IX, an endogenous intermediate in the heme synthesis pathway. The reason for the selective accumulation is not fully understood but believed to be related to the changes in metabolic activity of diseased compared to normal cells. Following photoactivation with red light (∼635 nm), targeted HSIL cells are selectively destroyed by generating ROS inducing apoptosis and necrosis as well as stimulating the systemic immune response with as ultimate goal histological regression and complete oncogenic HPV clearance while preserving the underlying cervical stroma (22, 28).

As topical administration was perceived to be inconvenient, the integrated light- and drug-delivery device Cevira^®^ was developed (22, 26, 31). The safety and efficacy of Cevira^®^ have been evaluated in a double-blind, placebo-controlled dose-finding Phase 2b study including 262 patients with CIN1/2 randomised to HAL 0.2%, 1% or 5% or placebo, permitting retreatment at 3 months if clinically indicated (26). Based on the outcomes of the Phase 2b, the HAL 5% dose was selected for further evaluation in the Phase 3 study. The HAL 5% dose had a favourable safety profile while being associated with the highest regression rate and oncogenic HPV clearance. However, efficacy could only be demonstrated in patients with CIN2, probably due to a high rate of spontaneous regression in the CIN1 population with most patients being HPV negative. As a result, the present Phase 3 study will only include HSIL patients (CIN2/3) ensuring efficacy can be reliably assessed. Due to the local and transient exposure to HAL, side effects were usually self-limiting, mainly including discharge, discomfort and spotting.

The currently available data indicate that Cevira^®^ is easy-to-use for gynaecologists and well-accepted by patients (31). The device results in no patient down-time as it is similar to using a tampon, patients may leave the gynaecologist office immediately after the application and can return to normal daily activities. Additionally, patients can easily remove the device themselves by pulling the string within 24 hours after application. The current Phase 3 study will further evaluate how the use of Cevira^®^ device is perceived by gynaecologists and patients.

In conclusion, Cevira^®^ holds potential to serve high unmet medical need for non-surgical, safe treatment options for patients with HSIL and cervical cancer. Due to its non-invasiveness, Cevira^®^ could be a promising alternative to excisional treatment for young women in reproductive age. Following the encouraging results from the Phase 2b study, the efficacy and safety of Cevira^®^ in patients with HSIL will be further evaluated in the presented APRICITY Phase 3 study currently recruiting patients in China and Europe for a multicentre, prospective, double-blind randomised controlled clinical trial.

## Data Availability

All data produced in the present study are available upon reasonable request to the authors

## Acknowledgments

The authors are grateful to Ismar Healthcare NV who provided medical writing assistance on behalf of Shanghai Yahong MediTech Co., Ltd. The authors would also like to thank Ctirad Mokraš for input regarding study design, outcome definition and ethical approval application.

## Author contributions

FC, LS, YZ, ZY, JH and PH contributed to the experimental design of the study. CD and ZN were involved in study design, outcome definition and ethical approval application. JL is the global PI of the study. PH is the European PI of the study. FC is the Chinese PI of the study. All PIs have contributed to the study protocol amendments. All authors have read, edited and approved the manuscript.

## Funding

This study and support in the process of manuscript development were funded by Shanghai Yahong MediTech Co., Ltd, Pudong, Shanghai, China.

## Conflict of interest

CD received consulting fees from MSD, GSK, Tesaro and Clovis Oncology and honoraria from MSD and GSK. LH is an employee of Asieris Pharmaceuticals (Shanghai) Co., Ltd. FC, ZN, LS, YZ, ZY, JL and PH have nothing to declare.

## Patient and public involvement

Patients and general public were not involved in the design, conduct, reporting or dissemination plans of the presented study. Patients will receive HPV and colposcopy test results from their provider, as part of routine practice.

## Notes

### Clinical Trial

NCT04484415

### Author Declarations

The study was approved by the ethics committee of the Peking Union Medical College Hospital and Hannover Medical University, Germany.

